# Differences in Mean Diffusivity values in the Corona Radiata between patients with Alzheimer’s Disease and Healthy Controls

**DOI:** 10.1101/2024.10.09.20222463

**Authors:** Daniel J. Asay, Naomi J. Hunsaker

## Abstract

Our understanding of Alzheimer’s disease (AD) is at an all time high. Despite the progress being made, AD is currently irreversible and the number of confirmed cases continues to grow world wide. There is a need for deeper understanding of the course of Alzheimer’s and the development of more sensitive diagnostic tools. In the present study, we performedDTI and TBSS analyses of the Corona Radiata (CR) and Uncinate Fasciculus (UF) in the brain. We extracted MD and FA values of 10 AD patients and compared them to 16 healthy controls using ENIGMA DTI protocol. We also analyzed the Mini-mental state examination (MMSE) scores of each subject. We predict that patients with AD will have lower FA scores and higher MD values compared to healthy controls. We also predict that higher MD scores will be predictive of lower MMSE scores. The results indicate a significant difference in MD values in the CR between groups, which indicates greater axonal necrosis in the CR in patients with AD. However, FA values were not different between groups and patients with higher MD values did not have lower MMSE scores.These results indicate that axonal necrosis in the CR does not have a significant effect on executive functioning as measured by the MMSE. However, these results provide support to the notion of using DTI methodologies as a means of diagnosing AD.

## Introduction

Our understanding of Alzheimer’s Disease (AD) is at an all-time high. Neuroscientists and psychologists are taking meaningful steps forward towards understanding AD at every level; from the behavioral and neurobiological level in humans, to the molecular and cellular level in animals (Grodzicki & Dziendzikowska, 2020; Lei et al., 2020). Despite this progress, AD is currently irreversible and treatments are largely aimed at alleviating symptoms (Velazquez, Anantharaman, Velazquez, & Lee, 2019). However, some researchers are optimistic about health outcomes for individuals who can be diagnosed at an earlier age when interventions may be more effective (Mueller et al., 2005). As life expectancy continues to rise, so does the number of individuals diagnosed with AD. As such, it is critical to understand the course of the disease as well as early signs of its onset. Previous studies indicate that the course of AD includes atrophy in all substructures of the corona radiata (CR) (Birdsill et al., 2014). The neurobiological function of CR includes somatosensory processing and executive control. As such, atrophy in the CR will likely lead to impaired executive functioning in patients with AD (Birdsill et al., 2014). Additionally, the Uncinate Fasciculus (UF) may be a site of atrophy in patients with AD. Atrophy in the UF has implications in loss of declarative memory function, which is one of the first indicators of cognitive decline in Alzheimer’s patients (Kiuchi et al., 2009; Papagno et al., 2011).

In the current study, we analyzed the CR and UF of patients with AD and healthy controls (HC) using MRI data from the EDSD database and performed a diffusion tensor imaging (DTI) analysis. DTI is a method of MRI data analysis used to map and measure the white matter tracts in a subjects brain that provides insight to the presence of neurodegeneration (Douaud et al., 2011). Additionally, we used tract-based spatial statistics (TBSS) to compare the eigenvalues of voxels in the CR and UF between the patients with AD and HC.

As a measure of brain atrophy, we used extracted FA and MD values from our DTI and TBSS analyses. As a measure of executive functioning, we used the mini-mental state examination test (Tombaugh, McDowell, Kristjansson, & Hubley, 1996). FA values of the CR and UF will provide insight into their micro-structural integrity (Tromp, 2016). Higher FA values indicate higher white-matter structural integrity, while lower FA values indicate a break down in white matter structure leading to a increased volume of cerebrospinal fluid (CSF) (Smith et al., 2006). MD values were extracted as a means of understanding the extent of axonal necrosis in the CR (Tromp, 2016). As such, higher MD values indicate greater axonal necrosis and therefore atrophy in white matter structural tracts.

We predict that there will be more atrophy in the CR and UF in patients with AD compared to HC. More specifically, we predict that patients with AD will have lower FA values and higher MD values in the CR and UF compared to HC (Mouton, Martin, Calhoun, Dal Forno, & Price, 1998). Additionally, we predict that higher MD values in the CR will be predictive of lower subject MMSE scores.

## Methods

### Participants

Participants included 17 females and 11 males from ages 53-85, with an average age of 68.5 and standard deviation of 8.17. Years of education ranged from 8-19 years, with an average of 13.73 years and standard deviation of 3.58. Additional demographic information is available in Table 1.

**Table 1.**
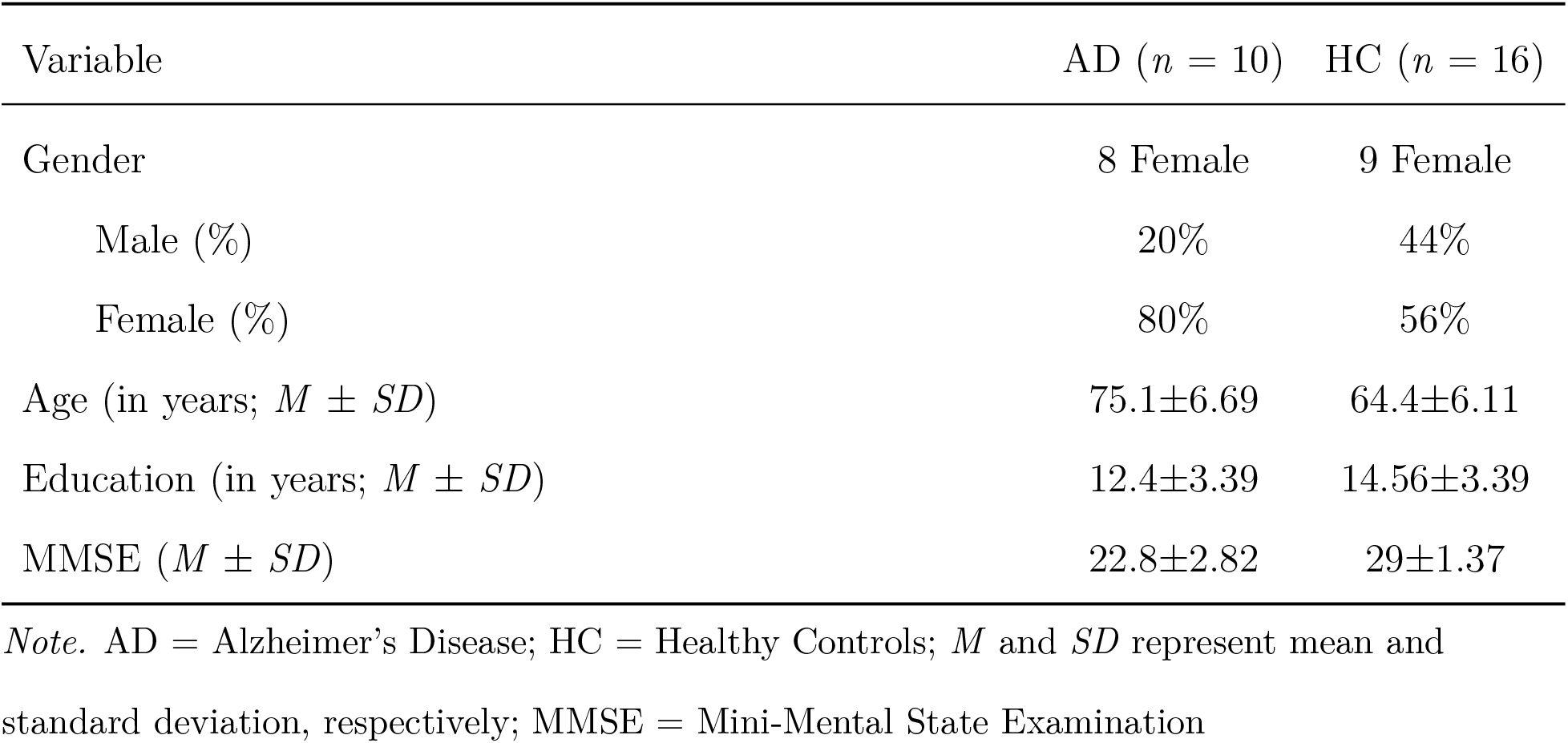
Characteristics of Study Population by Diagnosis.

### MRI Acquisition

Images were obtained from 1.5and 3.0 Tesla MRI scanners,including9 scanner models from 3 vendors. The number of gradient directions of the DTI scans varied between 6 and 61, and slice spacing varied between2 and 2.4 mm across the different scanners. In addition, anatomical 3D gradient echo T1-weighted MRI scans of approximately 1 mm3resolution were obtained from all scanners.

### Procedure

Computing resources were provided by the Brigham Young University Super Computer, which included access to 23,920 CPU cores and 90 TB of memory over 1,012 nodes run on Red Hat Enterprise Linux 7.6. Relevant information can be found at the following link https://rc.byu.edu/documentation/resources. Chris Rorden’s dcm2niix version v1.0.20190902 (Li, Morgan, Ashburner, Smith, & Rorden, 2016) was used convert neuroimaging data from the DICOM format to the NIfTI format. All data was organized in format consistent with version 1.0 (Gorgolewski et al., 2016). Acpcdetect version 2.0 was used to align the anterior and posterior commissure of each subject (Ardekani & Bachman, 2009). We used the convert3d tool for converting 3D images between common file formats (Yushkevich et al., 2006).

FSL version 6.0 was used for the brain extraction tool for deletion of non-brain tissue (Smith, 2002), eddy current correction to correct for eddy currents and movement during scanning (Andersson & Sotiropoulos, 2016), diffusion tensor fitting to fit a diffusion model at each voxel, and TBSS analysis for analyzing white matter data (Smith et al., 2006). We used ANTS version 2.2.0 N4BiasFieldCorrection for the remapping and smoothing of voxels (Tustison et al., 2010). ANTS version 2.2.0 was also used for registering subjects in standard space (Avants et al., 2011; Tustison & Avants, 2013). Individual subject FA maps were created using the ENIGMA DTI protocol (Kelly et al., 2018). The John Hopkins University White Matter Atlas was used as a reference for the construction of subject FA maps (Mori et al., 2008). Figure 1 shows the John Hopkins University White Matter Atlas.

**Figure 1.**
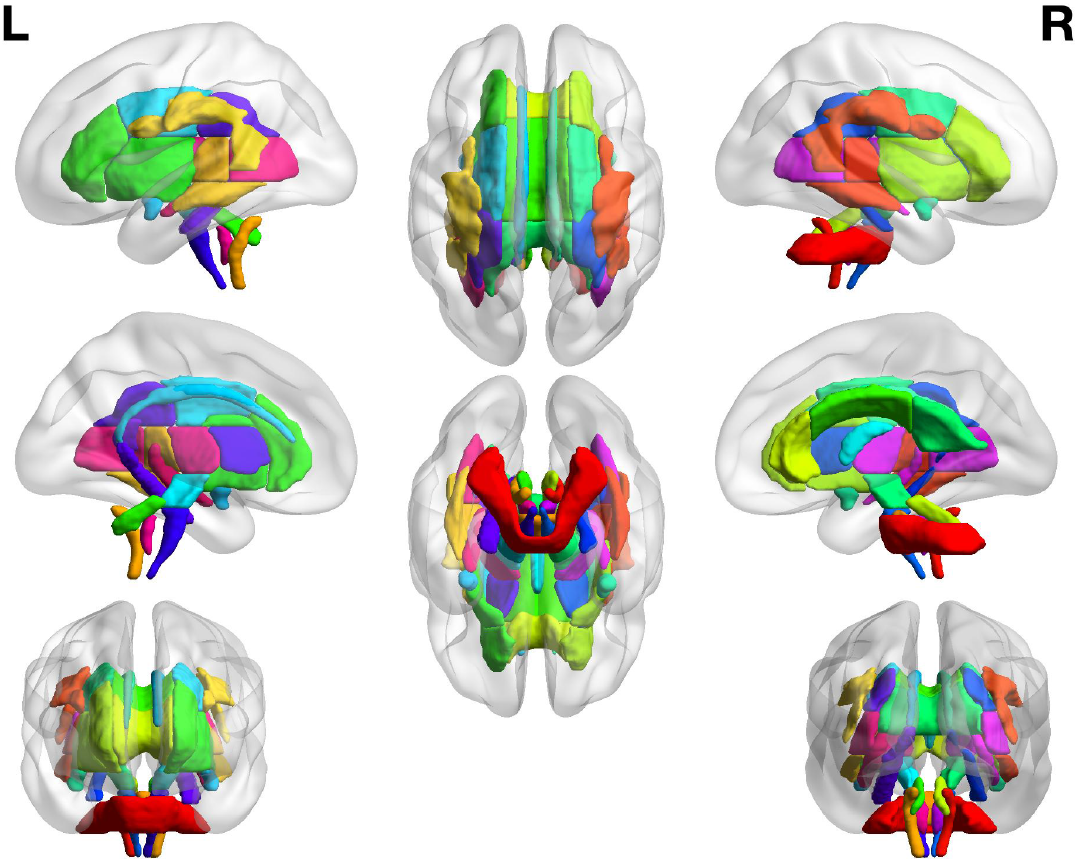
John Hopkins University White Matter Atlas

### Design and Analysis

A two-way ANOVA analysis will be conducted to measure differences in CR and UF atrophy and MMSE scores between AD patients and HC. Additionally, we will construct a linear model ANOVA to analyze the predictive power of MD values relative to executive functioning as represented by MMSE scores. All statistical analyes were performed in R version 3.6.3 (R Development Core Team, 2010).

## Results

### Summary Statistics

Exploratory and summary statistics can be found below in Table 2.

**Table 2.** Summary Statistics.

*Summary Statistics*
| Statistic | Group | CR-FA | CR-MD | UF-FA | UF-MD |
| --- | --- | --- | --- | --- | --- |
| Mean | AD | 0.4927 | 0.0007286 | 0.5827 | 0.0008474 |
| Standard Deviation | AD | 0.0357 | 3.07e-5 | 0.0569 | 0.0001196 |
| Mean | HC | 0.4918 | 0.0006765 | 0.6066 | 0.0007687 |
| Standard Deviation | HC | 0.0223 | 3.27e-5 | 0.0532 | 0.0001056 |

*Note.* AD = Alzheimer's Disease; HC = Healthy Control; CR = Corona Radiata; UF = Uncinate Fasciculus; FA = Fractional Anisotropy; MD = Mean Diffusivity

### ANOVA Results

We constructed a linear model for each of our variables of interest, which included FA and MD values in both the AD and HC group in the CR and UF. Based on that linear model, we ran a two-way ANOVA to test for differences in FA and MD values. All tests were run at the *α* = .05 significance level.

After conducting all ANOVA analyses, a significant difference between groups was detected in the CR MD values with p=.0014 and F=13.786. All other ANOVA analyses yielded p-values >.05. The results from the ANOVA analysis can be found in Table 3.

**Table 3.** ANOVA Results Table.

| Resp: Mean Diffusivity CR | DF | Sum of Squares | Mean Squares | F-Value | P-Value |
| --- | --- | --- | --- | --- | --- |
| Group | 1 | 1.57e-8 | 1.57e-8 | 13.786 | .00014 |
| MMSE Score | 1 | 3.8e-10 | 3.8e-10 | .355 | .5693 |
| Education | 1 | 1.904e-10 | 1.904e-10 | .169 | .6864 |
| Age | 1 | 4.134e-10 | 4.134e-10 | .364 | .5529 |
| Residuals | 20 | 2.27e-8 | 1.135e-9 | N/A | N/A |

*Note.* DF = Degrees of Freedom; Resp = Response Variable; N/A = Not Applicable;

An unpaired, two sided two-sample t-test was conducted to compare the means of the AD group and healthy control group MD values in CR. The results are as follows: t=3.8995, df=23, p-value=.0007218 with a 95% confidence interval of (2.45e-5, 7.98e-5). Group means for the AD and HC groups were .0007286438 and 0.0006765111 respectively.

### Brain Region Visualization

Group differences in MD values are shown in Figure 2. Figure 2 shows the corticospinal tract in the brain, which the CR forms a part of. Figure 2 was rendered in R version 3.6.3 using the JHU atlas (Mori et al., 2008; R Development Core Team, 2010).

**Figure 2.**
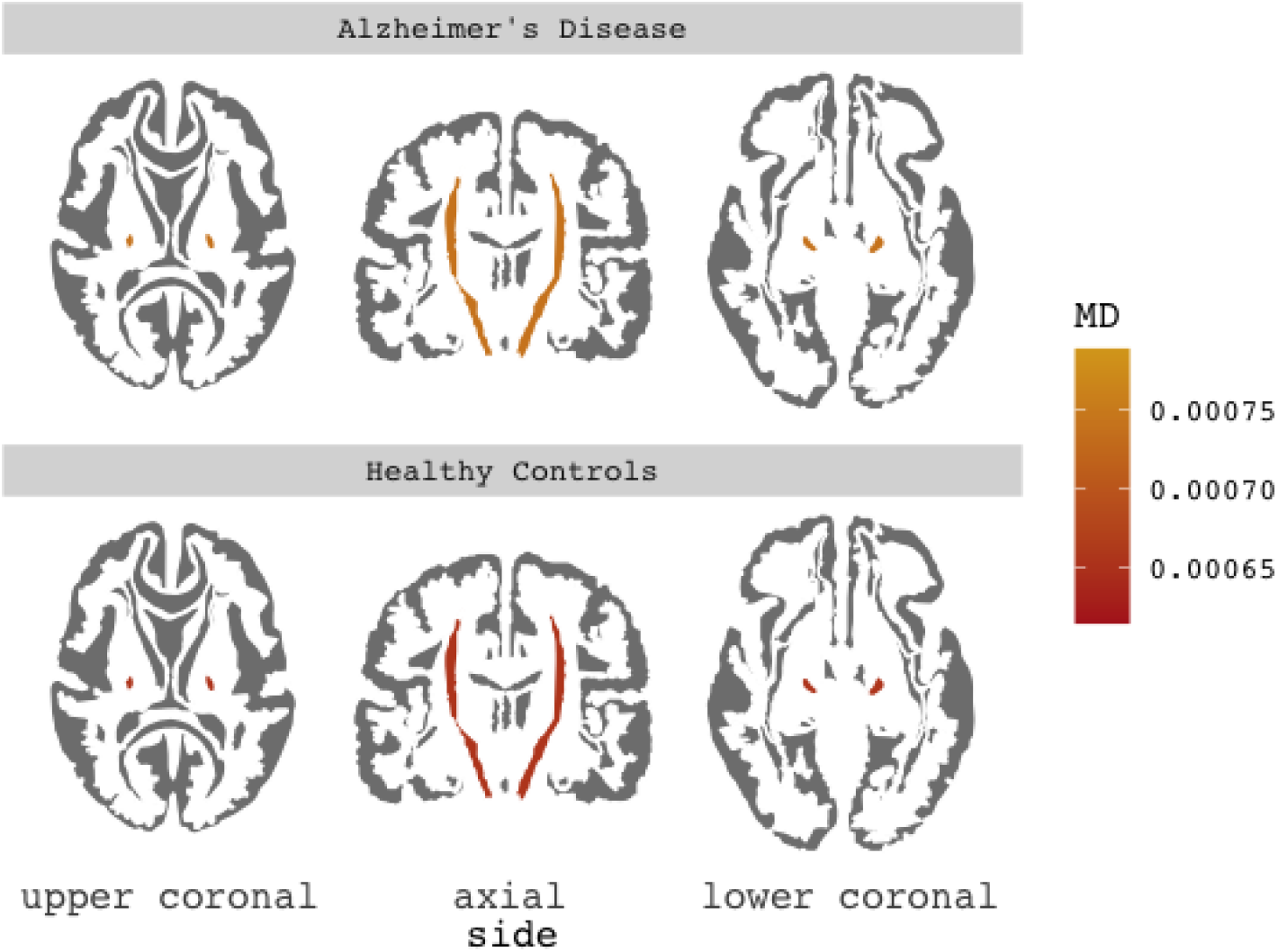
Group Differences in MD Values. Image Rendered from John Hopkins University White Matter Atlas.

## Discussion

The linear model ANOVA anlaysis indicated a significantly higher MD value in patients with AD, which indicates a higher degree of axonal necrosis in the CR (Tromp, 2016). As a result of increased axonal necrosis in the CR, we would expect to see a greater impairment in executive functioning as measured by the MMSE neuropsychological test (Tombaugh et al., 1996). However, our linear model ANOVA indicates that MMSE score is not a significant predictor of MD values in the CR. Additionally, when the linear model is built with MMSE score as the response variable, MD values are not a significant predictor of MMSE score. As such, our second hypothesis is partially supported; patients with AD have significantly higher MD values in the CR, however there is no support for patient CR MD values affecting cognition or vice versa. There was no evidence to support all other hypotheses.

### Practical Implications

From a clinical point of view, the findings from the present study may be useful. The MD values extracted from the DTI analysis of the CR successfully distinguished between patients with AD and HC. Therefore, extracting MD values from the CR may be a useful neurobiological diagnostic tool as well as in monitoring the progression of AD. As more DTI studies are done in AD, it is possible that an MD value threshold may be discovered for diagnostic purposes. Additionally, an MD value threshold could prove useful to clinicians when diagnosing AD versus mild cognitive impairment based on the amount of axonal necrosis.

The present study also has potential implications in the utility of the study of white matter tracts in dementia. Historically, grey matter has been of more interest to dementia researchers; however, this study provides support to the growing body of evidence that changes in white matter tracts are bio-markers for dementia (Chutinet & Rost, 2014). The continued use of DTI analysis is dementia can provide meaningful insights into the course of AD and, as explained previously, into the discovery of more sophisticated diagnostic tools.

### Strengths

The present study used the most cutting edge neuroimaging data analysis tools to tease apart the data. The use of the most updated versions of the various neuroimaging tools allowed for an intricate understanding of the white matter tracts in each subjects brain which led to robust results and conclusions. Additionally, the use of powerful analytic tools allows for high confidence in rendered results and provides greater ease of interpretation of results.

The subject of study in this project also serves as one of its fundamental strengths. Relatively few studies have analyzed the role of white matter fiber integrity in AD, and even fewer in the CR and UF specifically. The findings in the present study provide support to the growing body of white matter necrosis as a bio-marker for AD while also providing novel insights and data.

### Weaknesses

The sample size of this study is relatively low, with a total n of 26. Therefore, the generalizability and ecological validity of the results may have low reliability. Furthermore, a sample size of n < 30 teeters on the violation of normality in statistical analyses. In addition, the data used for the study was not directly collected by the authors. While information regarding scanning protocols were shared, it is impossible for the authors to be aware of possible inconsistencies and hiccups regarding neuroimaging data collection.

In future studies, results could be considered more reliable and robust with a greater sample size. With an n > 30, researchers would be able to draw more convincing conclusions and more accurately account for subject variability. Additionally, future studies should collect data on site, where possible. Doing so would allow for sufficient knowledge of aberrant data collection events and facilitate correction of those events.

### Future Directions

Based on the results found here, future studies should focus on the use of extracting MD values from the CR as a means of diagnosing AD. A methodology diagnostic study of DTI and MD values in the CR has implications for catching AD earlier in life, perhaps even before official neuropsychological diagnosis, which would allow for earlier implementation of treatment to curb the detrimental cognitive effects of AD. Future studies could also focus on conducting longitudinal studies that compare white-matter tract differences in patients with early-onset AD versus late-onset AD. Such studies would provide insight into the neurobiological course of AD and the possible differences in said course in early-onset and late-onset patients.

## Conclusion

The present study provides additional evidence to the body of literature that axonal necrosis occurs in the CR in patients with AD. Additionally, axonal necrosis in the CR occurs at a statistically significant rate when compared to age-matched controls. However, axonal necrosis in the CR seems to have little to no effect on executive functioning as measured by the MMSE. Furthermore, years of education and age were not significant predictors of FA and MD values or MMSE score. The results found here serve as another piece of evidence for the use of DTI analysis as a diagnostic tool for AD and for deeper understanding of the neurobiological course of AD.

## Data Availability

All data used is available on the EDSD database.

https://www.researchgate.net/deref/https://neugrid4you.eu

